# Consistency of Serial CSF alpha-Synuclein Seed Amplification Assay Results in the Parkinson’s Progression Marker Initiative

**DOI:** 10.64898/2026.04.01.26349969

**Authors:** David G. Coughlin, Caroline Gochanour, Jie Yin, Luis Concha-Marambio, Carly Farris, Yihua Ma, David-Erick Lafontant, Edwin Jabbari, Tanya Simuni, Ken Marek, Thomas F. Tropea, the Parkinson’s Progression Marker Initiative

**Affiliations:** Department of Neurosciences, University of California San Diego, La Jolla CA; Department of Biostatistics, University of Iowa, Iowa City IA; Research and Development Department, Amprion, San Diego, CA; Department of Clinical and Movement Neurosciences, UCL Queen Square Institute of Neurology, London, UK; Department of Neurology, Northwestern University, Chicago IL; Institute for Neurodegenerative Disease, New Haven CT; Department of Neurology, University of Pennsylvania, Philadelphia PA

## Abstract

Studies reporting alpha-synuclein seed amplification assay (aSyn-SAA) results are often cross-sectional. Here we investigated the intra-individual consistency of aSyn-SAA results over time from participants in the Parkinson’s Progression Marker Initiative (PPMI). A total of 1238 participants had >1 CSF aSyn-SAA result for analysis (Parkinson’s disease [PD]=633, prodromal =563, healthy control [HC]=42) which were collected over a median (min, max) of 2.0 (0.4, 11.4) years. Emphasis was placed on evaluating consistency in less common results such as aSyn-SAA- PD participants, aSyn-SAA+ HC and conversion rates from aSyn-SAA negative to positive results prodromal participants. Of aSyn-SAA+ PD participants, 96% (474/493, 95%CI 94-98%) remained positive in subsequent samples, and 92% (116/126, 95%CI 86-96%) of aSyn-SAA- PD participants remained negative. 99% (303/307, 95%CI 97-99%) of aSyn-SAA+ prodromal participants remained positive, and 95% (234/247, 95%CI 91-97%) of aSyn-SAA- prodromal participants remained negative. 89% (16/18, 95%CI 67-97%) of aSyn-SAA+ HC participants remained positive, and 87% (20/23, 95%CI 68-95%) of aSyn-SAA- HC participants remained negative. These results confirm a high consistency of aSyn-SAA results over time, even in less expected results.

## Introduction

Alpha-synuclein seed amplification assays (aSyn-SAA) measured in the cerebrospinal fluid (CSF) have high sensitivity and specificity in identifying aSyn neuropathology in people with Parkinson’s disease (PD), dementia with Lewy bodies, multiple systems atrophy, prodromal states like idiopathic REM sleep behavior disorder (iRBD) and hyposmia(1–4). However, the intra-individual consistency of aSyn-SAA outcomes in serially collected samples has only been investigated in dementia with Lewy bodies, Alzheimer’s disease, and in participants with genetic variants placing them at higher risk for developing PD (5–7). To date, longitudinal consistency has not been reported in PD or in large cohorts of participants in prodromal states without genetic variants. As CSF aSyn-SAA becomes more widely used in clinical trials, it would also be important to understand the consistency or assay results over time. Here we investigate the longitudinal consistency of aSyn-SAA results in individuals from different cohorts in the Parkinson’s Progression Marker Initiative (PPMI) study with three goals: 1) to examine the consistency of aSyn-SAA results over time in the setting of typical results including aSyn-SAA+ PD and aSyn-SAA- healthy participants, 2) to examine the consistency of aSyn-SAA results over time in the setting of infrequent results including aSyn-SAA- PD participants and aSyn-SAA+ healthy participants and 3) to examine aSyn-SAA status in prodromal states where changes may represent disease progression.

## Methods

### Cohort Selection

The PPMI study is an international observational study across academic neurology centers in multiple countries, aimed at identifying clinical and biological markers of Parkinson’s disease heterogeneity and progression. PPMI is registered at ClinicalTrials.gov (NCT01141023), with IRB approval at each site and written informed consent from participants. Full inclusion criteria and study design details are available on the PPMI website at ppmi-info.org.

Participants with one or more CSF aSyn-SAA result were available from three cohorts:

- **Parkinson’s Disease (PD)**: For participants without pathogenic genetic variants (i.e. sporadic PD [sPD]), inclusion required diagnosis within 2 years of enrollment, no prior PD medication, Hoehn & Yahr stage 1–2(8), abnormal DAT-SPECT, and at least two cardinal motor signs (resting tremor or bradykinesia required). Participants with pathogenic genetic variants (e.g. *LRRK2, GBA1, SNCA, PRKN*, etc.) could have started PD medications and have disease duration of up to 7 years at enrollment.
- **Prodromal**: Participants had hyposmia and/or evidence of RBD (either polysomnogram proven iRBD or reports of dream enactment behavior) or carried pathogenic genetic variants, (i.e. non-manifesting carriers [NMC]). Prodromal participants did not have clinical diagnoses of PD.
- **Healthy control (HC)**: HC participants had no parkinsonism, dementia, or major neurological/psychiatric disease.

### CSF aSyn-SAA

All aSyn-SAA testing was performed by Amprion (San Diego, CA) by staff members who were blinded to clinical diagnoses and cohort assignments of participants. Over the course of the PPMI study, three versions of the CSF aSyn-SAA have been used: 150-, 24-, and 35-hour (h) versions. All three versions give a positive, negative, or inconclusive result based on triplicate runs surpassing a fluorescence threshold. The 24h and 35h assays additionally discriminate between Type-I (typically seen in PD and dementia with Lewy bodies) and Type-II (typically seen in multiple systems atrophy) seeding activity as described previously (4,9). Individual participants could have had different versions of the CSF aSyn-SAA assay performed at different time points. If a participant had different versions of the assay run on samples collected at a single visit, the result from the most recently run assay was used for analysis.

## Statistical Analysis

Summary statistics, including counts for aSyn-SAA results, and medians, minimum, and maximum for the time intervals between assays are provided. To assess consistency rates, point estimates and 95% Wilson confidence intervals were calculated. All analysis used SAS v9.4 (SAS Institute Inc., Cary, NC, USA; sas.com; RRID:SCR_008567).

## Results

One thousand two hundred thirty-eight participants of the PPMI study had more than one CSF aSyn-SAA results available for analysis (PD n=633, prodromal n=563, HC n=42). Among these participants, there were 2738 total CSF aSyn-SAA results available with a median (min, max) number of 2 (2, 7) samples per participant. Samples were collected over a median (min, max) time of 2.0 (0.4, 11.4) years (**Table 1** and **Supplemental Material**). This included 680 samples with results from the 150h assay and 2,058 with results from the 24h or 35h assays. **Table 1** includes initial CSF aSyn-SAA results for PPMI participants with one aSyn-SAA result and then for the subset of participants with >1 result by cohort and subgroup. The latter were enriched with aSyn-SAA- PD and aSyn-SAA+ HC participants who were preferentially selected to have serial testing to understand result consistency in these groups; however, the percent of aSyn-SAA+ PD and aSyn-SAA- HC participants is therefore reduced compared to the entire PPMI cohort due to this approach.

**Table 1.** CSF aSyn-SAA longitudinal results across cohort and subgroup.

| Cohort | Subgroup | All Participants with CSF SAA Results |  |  |  |  | Participants with N > 1 Visits with CSF SAA Results |  |  |  |  |
| --- | --- | --- | --- | --- | --- | --- | --- | --- | --- | --- | --- |
|  |  | N any SAA results | N first visit pos./type I | N first visit negative | N first visit type II | N first visit inconclusive | N with N>1 result | N first visit pos./type I | N first visit negative | N first visit type II | N first visit inconclusive |
| PD | **Total** | **1389** | **1171 (84%)** | **195 (14%)** | **15 (1%)** | **8 (<1%)** | **633** | **493 (78%)** | **126 (20%)** | **9 (1%)** | **5 (<1%)** |
|  | sPD | 1032 | 930 (90%) | 85 (8%) | 11 (1%) | 6 (<1%) | 444 | 368 (83%) | 65 (15%) | 7 (2%) | 4 (<1%) |
|  | LRRK2 PD | 171 | 110 (64%) | 60 (35%) | 1 (<1%) | 0 | 125 | 76 (61%) | 48 (38%) | 1 (<1%) | 0 |
|  | GBA PD | 90 | 82 (91%) | 6 (7%) | 1 (1%) | 1 (1%) | 42 | 35 (83%) | 5 (12%) | 1 (2%) | 1 (2%) |
|  | Other Genetic PD | 54 | 38 (70%) | 16 (30%) | 0 | 0 | 22 | 14 (64%) | 8 (36%) | 0 | 0 |
|  | PD Normosmic | 42 | 11 (26%) | 28 (67%) | 2 (5%) | 1 (2%) | 0 | 0 | 0 | 0 | 0 |
| Prodromal | **Total** | **2471** | **1563 (63%)** | **891 (36%)** | **7 (<1%)** | **10 (<1%)** | **563** | **307 (55%)** | **247 (44%)** | **6 (1%)** | **3 (<1%)** |
|  | PSG+ iRBD & Hyposmia | 217 | 204 (94%) | 11 (5%) | 1 (<1%) | 1 (<1%) | 63 | 60 (95%) | 3 (5%) | 0 | 0 |
|  | PSG+ iRBD & Normosmia | 109 | 52 (48%) | 56 (51%) | 1 (<1%) | 0 | 38 | 12 (32%) | 25 (66%) | 1 (3%) | 0 |
|  | Self-Reported DEB & Hyposmia | 699 | 579 (83%) | 114 (16%) | 3 (<1%) | 3 (<1%) | 139 | 102 (73%) | 34 (24%) | 3 (2%) | 0 |
|  | Hyposmia Only | 966 | 641 (66%) | 319 (33%) | 1 (<1%) | 5 (<1%) | 261 | 121 (46%) | 137 (52%) | 1 (<1%) | 2 (<1%) |
|  | Genetic NMCs | 433 | 48 (11%) | 383 (88%) | 1 (<1%) | 1 (<1%) | 58 | 10 (17%) | 46 (79%) | 1 (2%) | 1 (2%) |
|  | Other Prodromal | 47 | 39 (83%) | 8 (17%) | 0 | 0 | 4 | 2 (50%) | 2 (50%) | 0 | 0 |
| HC | **Total** | **323** | **25 (8%)** | **297 (92%)** | **0** | **1 (<1%)** | **42** | **18 (43%)** | **23 (55%)** | **0** | **1 (2%)** |

Abbreviations: sPD: sporadic PD. LRRK2: Leucine rich repeat kinase 2. GBA: glucosylceramidase beta 1. PSG: polysomnogram. iRBD: idiopathic REM sleep behavior disorder. DEB: dream enactment behavior. NMC: non-manifesting carrier. HC: healthy control.

Of the overall PPMI cohort with at least one CSF aSyn-SAA result, 90% (930/1032) of sporadic PD (sPD) participants, 63% (1563/2471) of prodromal participants, and 8% (25/323) of HC participants had positive CSF aSyn-SAA testing at their initial testing visits. Of participants with >1 CSF aSyn-SAA result, 83% (368/444) of sPD participants, 61% (76/125) of *LRRK2*-PD, 83% (35/42) of *GBA*-PD participants had positive (for the 150h assay) or Type-I seeding patterns (for 24h or 35h assays) at their initial testing visits (**Table 1** with clinical characteristics in **Supplemental Material**). 95% (60/63) of participants with iRBD and hyposmia (≤15^th^ percentile of age and sex expected performance on the University of Pennsylvania Smell Identification test) had positive initial CSF aSyn-SAA results, as did 32% (12/38) of normosmic participants with iRBD, 73% (102/139) of participants with a clinical history dream enactment behavior with hyposmia, and 46% (121/261) of participants with hyposmia but no reported history of dream enactment. In the overall PPMI cohort, HC participants have a positive CSF aSyn-SAA frequency of 8%. For sake of this study, serial testing was preferentially obtained in these individuals resulting in a cohort of HC participants with >1 CSF aSyn-SAA result that had a 43% (18/42) positive CSF aSyn-SAA at baseline. Throughout the cohorts, there were small numbers of participants with Type-II seeding or inconclusive results (1% and <1% respectively).

Of PD participants with initial positive or Type-I seeding results, 96% (474/493, 95%CI 94-98%) continued to have positive or Type-I results on subsequent assessments. Of those with negative initial results, 92% (116/126, 95%CI 86-96%) continued to have negative results.

In prodromal participants with initial positive or Type-I seeding, 99% (303/307, 95%CI 97-99%) remained positive or with positive Type-I seeding and 95% (234/247, 95%CI 91-97%) of those with negative results remained negative in subsequent samples over a median (min, max) of 2.1 (0.4, 5.2) years and 2.0 (0.6, 8.9) years of observation respectively.

In HC participants with initial positive or positive Type-I seeding, 89% (16/18, 95%CI 67-97%) remained positive or with Type-I seeding and 87% (20/23, 95%CI 68-95%) with initial negative results remained negative subsequently (**Table 2**).

**Table 2.**
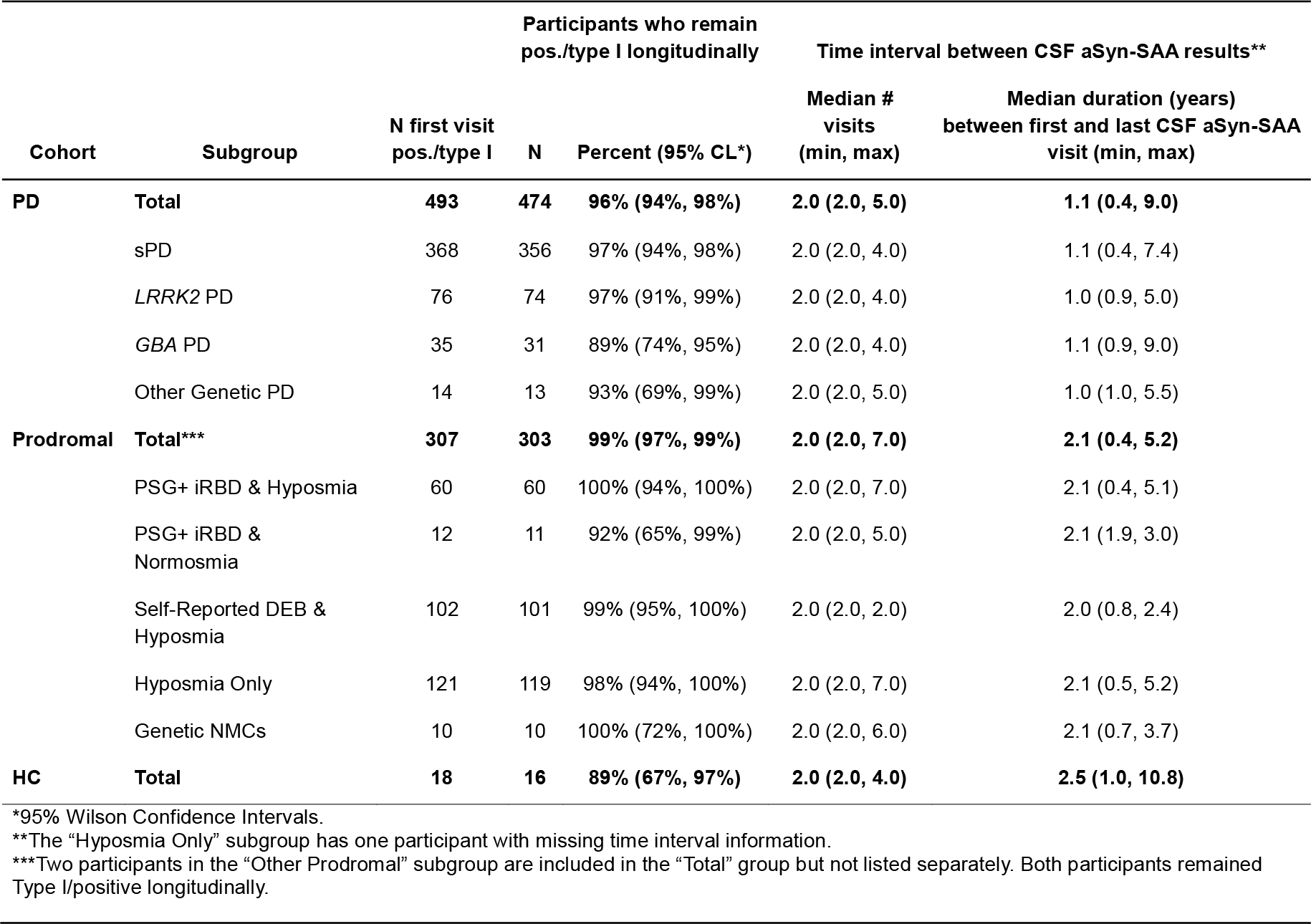
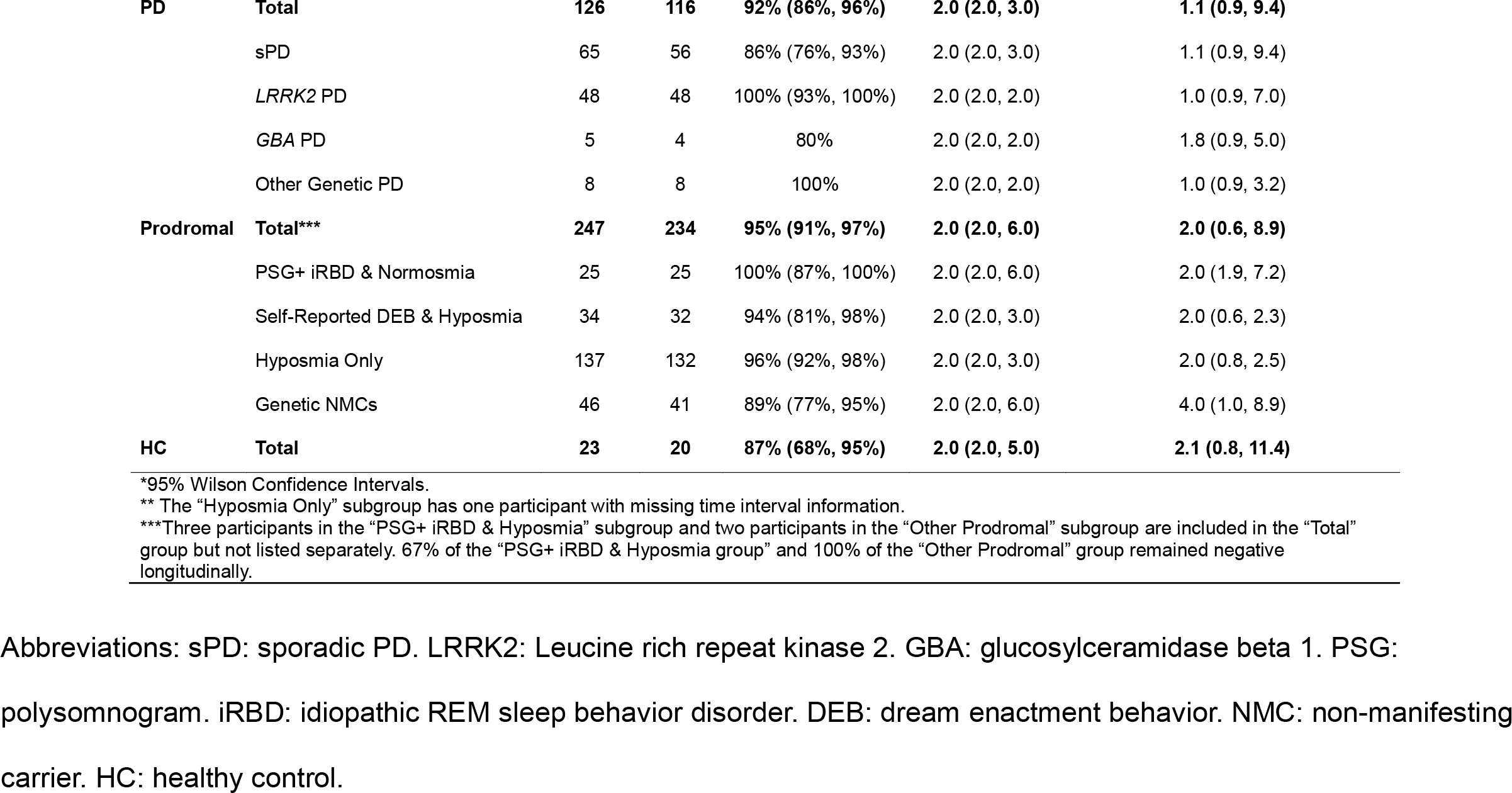
Longitudinal consistency of CSF aSyn-SAA results among those with N > 1 visits.

Two percent (n=11) of participants in the prodromal group with initial negative tests subsequently developed positive CSF aSyn-SAA results which occurred at a median (min, max) time of 2.4 (1.1, 5.5) years from initial assessment, potentially reflecting a progression of pathological processes over this time period. Because the duration of follow-up was longer for the prodromal cohort than for other cohorts, results from an analysis restricted to 2 years of follow-up testing is added to the **Supplemental material**. Numbers of participants with inconsistent longitudinal CSF aSyn-SAA results or Type-II seeding results were too few to justify statistical comparisons of clinical characteristics (**Table 1** and **Supplemental Material**).

## Discussion

In this study, we sought to understand how CSF aSyn-SAA results change over time using the large number of CSF samples from deeply phenotyped participants in PPMI. We noted very high rates of consistent results in this study, indicating limited changes and high reliability over the course of this study. This study is of importance as it provides insight into the consistency of aSyn-SAA over time in PD, HC, and prodromal participants. Specifically, the results argue against the need for frequent repeated measures over time to confirm results specifically when the results are biologically expected (CSF aSyn-SAA+ PD, CSF aSyn-SAA- HC). Results were consistent even in cases with less common situations such as aSyn-SAA- PD participants or aSyn-SAA+ HC participants. The overwhelming majority of CSF aSyn-SAA- prodromal participants remained such over the duration of ascertainment. PPMI is planning to continue testing of prodromal participants every two years to further investigate rates of change in aSyn-SAA- prodromal participants.

Highly consistent longitudinal positive and negative CSF aSyn-SAA results have been reported in cohorts of clinically diagnosed dementia with Lewy bodies, mild cognitive impairment and Alzheimer’s disease (6,7). Additionally, in a recent study primarily of participants with pathogenic *LRRK2* and *GBA* variants without clinical PD diagnoses (NMC), low numbers of participants developed positive tests over the course of their two-year study(5). This study included data from a large number of PD participants as well as non-manifesting non carrier prodromal participants. We also noted high degrees of consistency in both positive and negative CSF aSyn-SAA results in these and all cohorts studied here.

There are limitations of this study. It is possible that prodromal participants could develop aSyn-SAA positivity in the future. Further data collection and analyses are ongoing in this regard. Additionally, three versions of the Amprion aSyn-SAA were used in this study, which was necessary as the assay has been improved over the course of the PPMI study. We noted that testing with the 24h or 35h version of the assay was highly consistent with results from the 150h assay. 523 participants had results from different assay versions (150h and 24h/35h) run at different time points. 96% (502/523) of such participants had consistent results even in this scenario (432/448 (96%) consistently positive, and 70/75 (93%) consistently negative). Moreover, if 150hr assay results were excluded, 98% (365/374) participants that were initially type I positive remained type I longitudinally and 90% (304/336) participants that were initially negative remained negative over time. It remains possible that by using multiple versions of the assay, the estimates of consistency provided here may be lower than they would be otherwise.

Overall, however, these results indicate that a single CSF aSyn-SAA test is likely sufficient in most cases to characterize a participant’s aSyn biomarker profile for at least a two-year period.

## Data Availability

Data used in the preparation of this article were obtained on January 5th, 2026 from the Parkinson's Progression Markers Initiative (PPMI) database (www.ppmi-info.org/access-data-specimens/download-data), RRID:SCR 006431. For up-to-date information on the study, visit www.ppmi-info.org. This analysis was conducted by the PPMI Statistics Core and used actual dates of activity for participants, a restricted data element not available to public users of PPMI data. This analysis used aSyn-SAA results for participants of the Prodromal Cohort, obtained from PPMI upon request after approval by the PPMI Data Access Committee. Statistical analysis codes used to perform the analyses in this article are shared on Zenodo [10.5281/zenodo.18894423].

## Data Availability

Data used in the preparation of this article were obtained on January 5^th^, 2026 from the Parkinson’s Progression Markers Initiative (PPMI) database (www.ppmi-info.org/access-data-specimens/download-data), RRID:SCR 006431. For up-to-date information on the study, visit www.ppmi-info.org. This analysis was conducted by the PPMI Statistics Core and used actual dates of activity for participants, a restricted data element not available to public users of PPMI data. This analysis used aSyn-SAA results for participants of the Prodromal Cohort, obtained from PPMI upon request after approval by the PPMI Data Access Committee. Statistical analysis codes used to perform the analyses in this article are shared on Zenodo [10.5281/zenodo.18894423].

Protocol information for The Parkinson’s Progression Markers Initiative (PPMI) Clinical - Establishing a Deeply Phenotyped PD Cohort. can be found on protocols.io or by following this link: https://dx.doi.org/10.17504/protocols.io.n92ldmw6ol5b/v2.

## Notes

Funding: DC is supported by NINDS (NS120038), EJ is supported by a Medical Research Council Clinician Scientist Fellowship (UKRI1388).

### Competing Interest Statement

David Coughlin has recieved research funding from the the NIH and Michael J Fox Foundation (MJFF). He has received in-kind support from Amprion and CND Life sciences.
Luis Concha-Marambio is an Amprion employee of Amprion, a biotech company dedicated to the commercialization of seed amplification assay technologies, and is an inventor of several patents related to seed amplification assay technologies, which have been assigned to Amprion
Carly Farris is an employee of Amprion, a biotech company dedicated to the commercialization of seed amplification assay technologies, and is an inventor of several patents related to seed amplification assay technologies, which have been assigned to Amprion
Yihua Ma is an Amprion employee of Amprion, a biotech company dedicated to the commercialization of seed amplification assay technologies and is an inventor of several patents related to seed amplification assay technologies, which have been assigned to Amprion
Tanya Simuni, MD has served as a consultant for Blue Rock Therapeutics, Centessa, Critical Path for Parkinson's Consortium (CPP), MJFF, Prevail/Lilly, Roche/Genentech, Ventus, Sinopia, Takeda, Vanqua Bio, Ventus, Ventyx, and VIMA Tx. Dr. Simuni has equity in Sinopia and has served on the ad board for AskBio, Biohaven, Booster, GAIN, Janssen, Neuron23, Novartis, Parkinson Study Group, Prevail/Lilly, and Roche/Genentech. Dr. Simuni has served as a member of the scientific advisory board of Koneksa and UCB. Dr. Simuni has received research funding from Neuroderm, Prevail, Roche NINDS, MJFF, and Parkinson's Foundation.
Thomas Tropea declares no conflicts of interest.
Caroline Gochanour declares
Jie Yin declares
Edwin Jabbari declares no conflicts of interest
Ken Marek declares support to his institution (Institute for Neurodegenerative Disorders) from The Michael J Fox Foundation. Consultant for the Michael J Fox Foundation, Mitro, Roche, BMS, GAIN, Calico, Sanofi, Teva, Biohaven, GEHC, Lilly, ABLi, and Prothena.

### Funding Statement

DC is supported by NINDS (NS120038), EJ is supported by a Medical Research Council Clinician Scientist Fellowship (UKRI1388).
PPMI - a public-private partnership - is funded by the Michael J. Fox Foundation for Parkinson's Research and funding partners, including AbbVie, Alamar Biosciences, Aligning Science Across Parkinson's (ASAP), Arrowhead Pharma, Arvinas, AskBio, BIAL, BioArctic, Biohaven, BlueRock Therapeutics, Bristol Myers Squibb, Calico Labs, Capsida Biotherapeutics, Critical Path Institute, DaCapo Brainscience, Denali, Edmond J. Safra Foundation, Eli Lilly, Gain Therapeutics, GE Healthcare, Genentech, GSK, Insitro, Johnson & Johnson Innovative Medicine, Lundbeck, Merck, Neumora, Neuron23, Novarti, Olink, Regeneron, Roche, Sanofi, Tenvie, UCB, Vanqua Bio, Voyager Therapeutics, The Weston Family Foundation

### Author Declarations

PPMI is registered at ClinicalTrials.gov (NCT01141023), with IRB approval at each site and written informed consent from participants. Full inclusion criteria and study design details are available on the PPMI website at ppmi-info.org.

