## Supplemental Material for "Consistency of Serial CSF alpha-Synuclein Seed Amplification Assay Results in the Parkinson’s Progression Marker Initiative"

**Supplementary Material**

Date of download from LONI: 05JAN2026

**Contents:**

Supplementary Figures 1A-C: Timing of CSF aSyn-SAA testing in PD, Prodromal, and HC Cohorts for those with >1 result

Supplementary Tables 1. Longitudinal CSF aSyn-SAA results, demographics, and characteristics across cohort and subgroup with clinical characteristics

Supplementary Table 2. Longitudinal consistency of CSF aSyn-SAA results among those with N > 1 visits with clinical characteristics

Supplementary Tables 3A-B. CSF aSyn-SAA longitudinal consistency – capped at Year 2

PPMI Authorship Information

Supplementary Figure 1A: Timing of CSF aSyn-SAA testing for PD cohort for those with >1 result

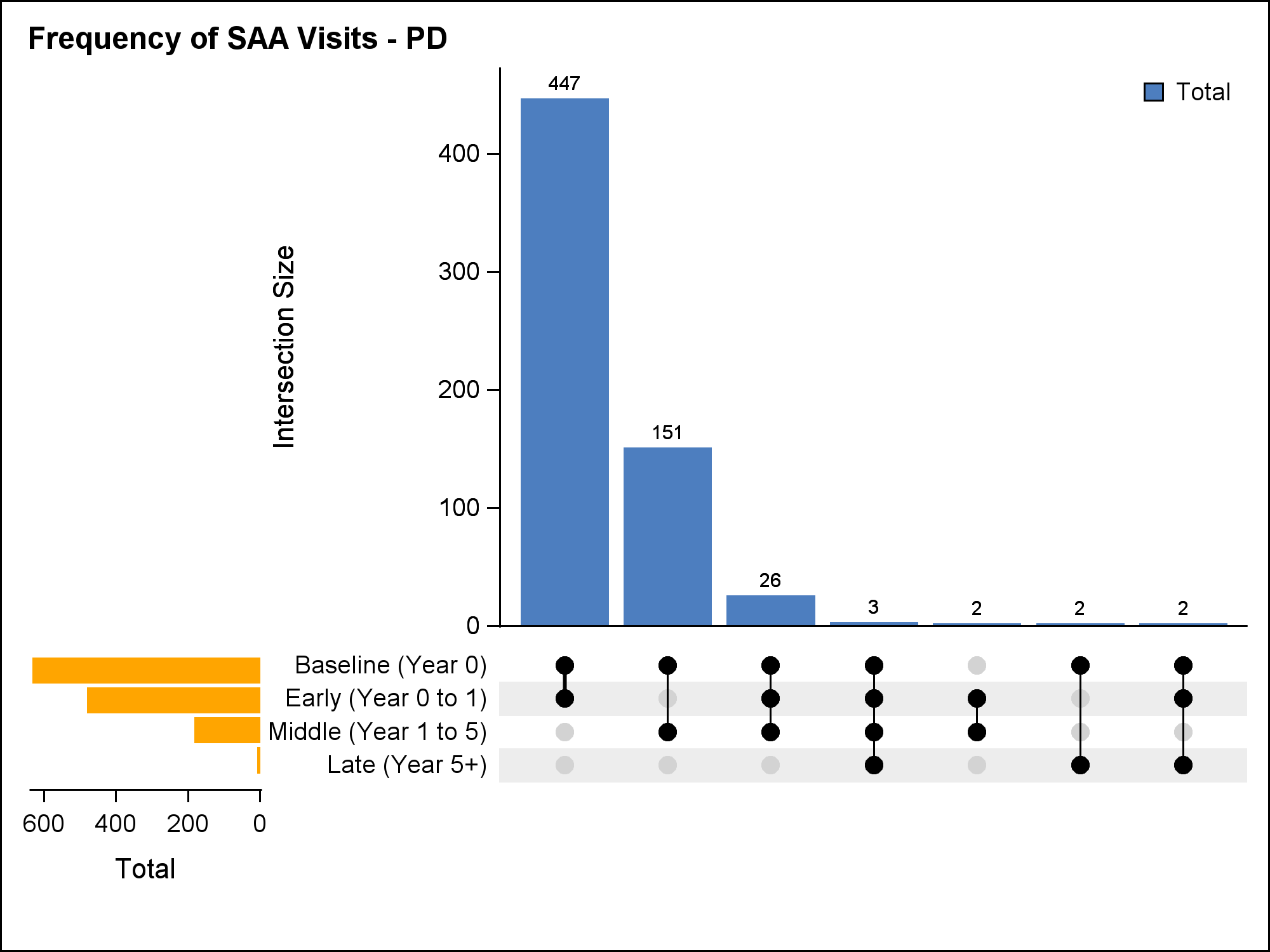

Supplementary Figure 1A: Timing of CSF aSyn-SAA testing for Prodromal cohort for those with >1 result

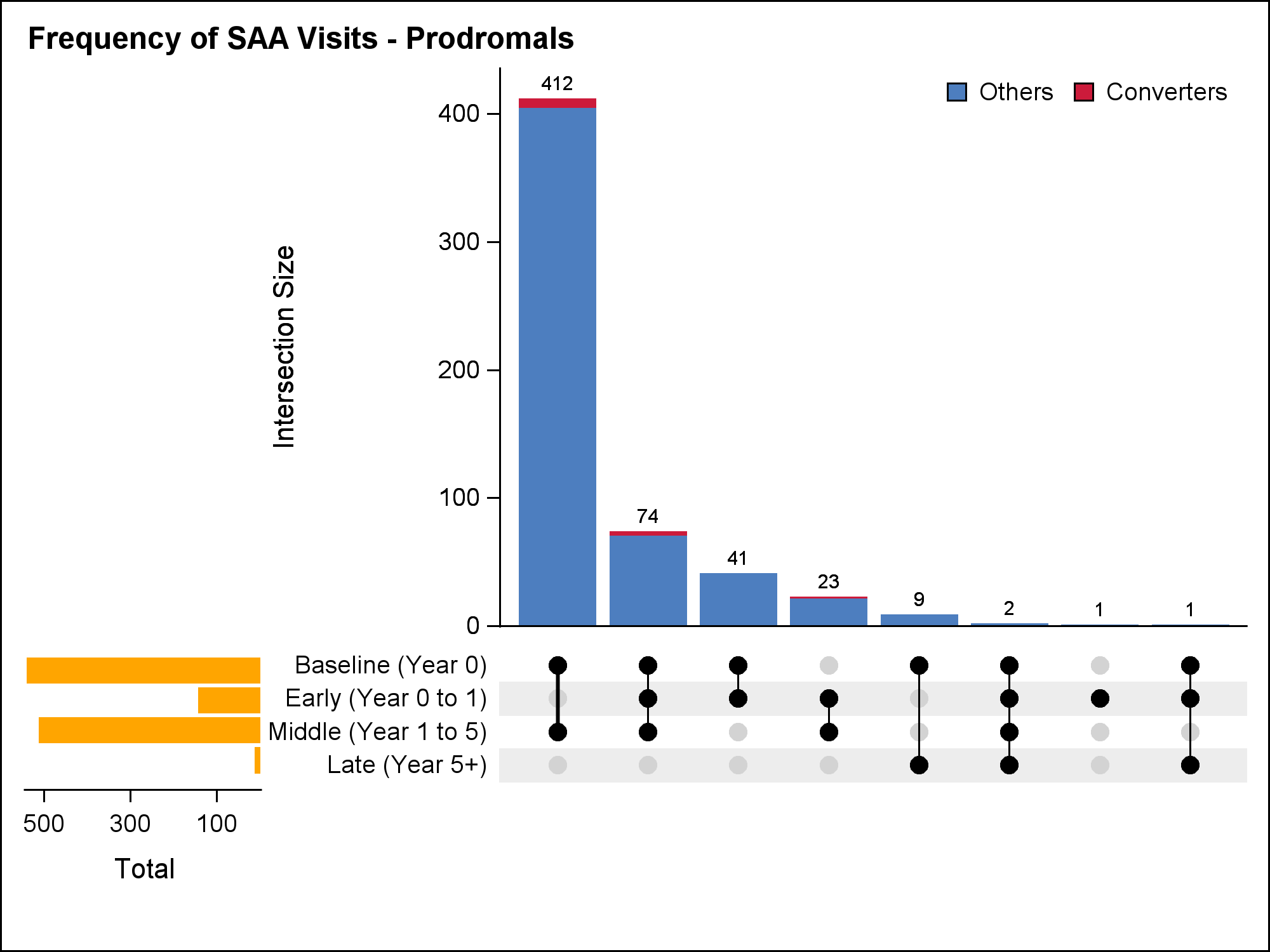

“Converters” are defined as participants that were initially SAA – and changed to SAA + longitudinally.

7/412 with Baseline and Middle results, 3/74 with Baseline, Early, and Middle results, and 1/23 with Early and Middle results are converters.

Supplementary Figure 1C: Timing of CSF aSyn-SAA testing for HC cohort for those with >1 result

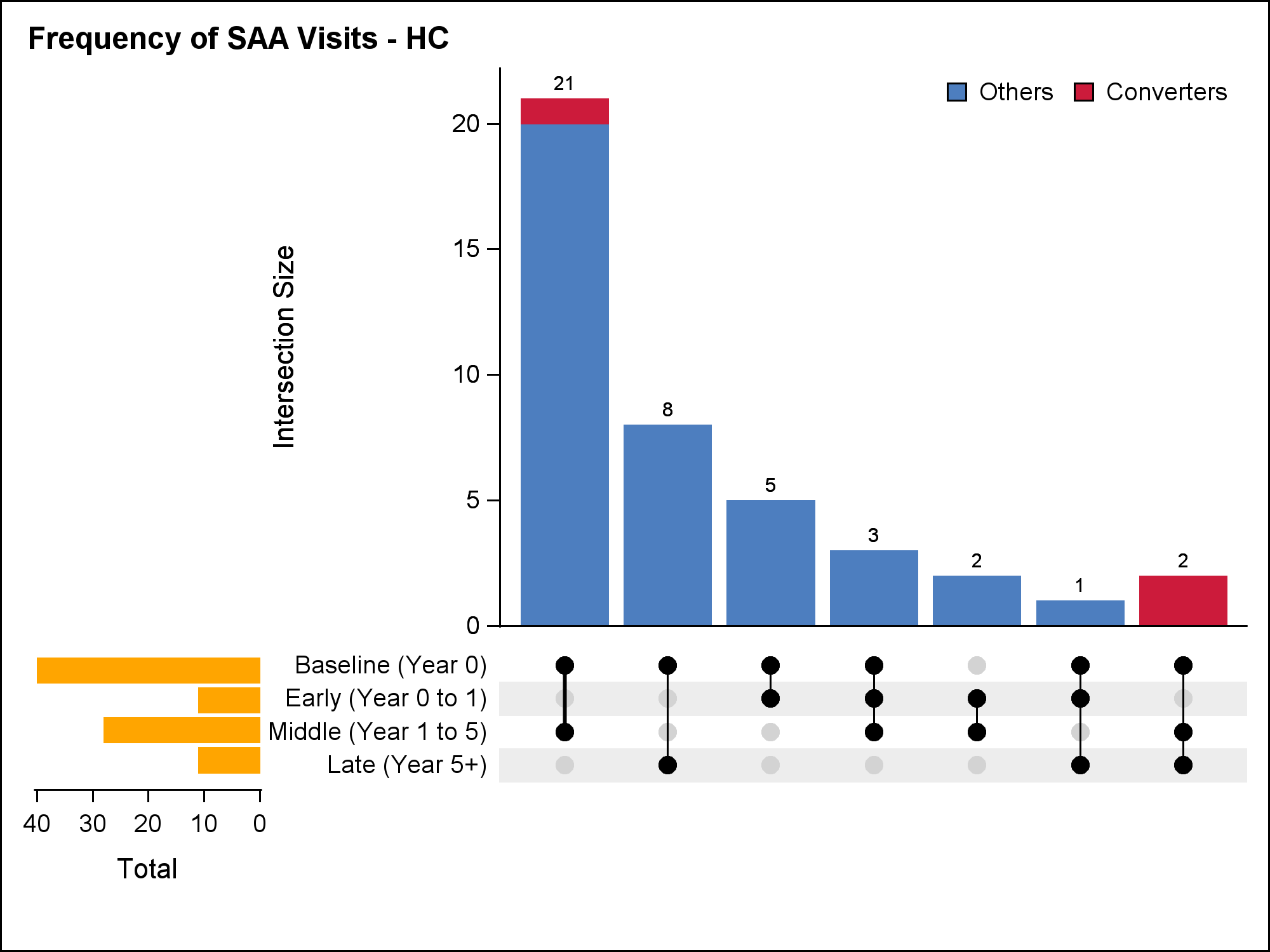

“Converters” are defined as participants that were initially SAA – and changed to SAA + longitudinally.

1/21 with Baseline and Middle results and 2/2 with Baseline, Middle and Late results are converters.

Supplementary Table 1. Longitudinal CSF aSyn-SAA results, demographics, and characteristics across cohort and subgroup

|  | | **Participants with N>1 Visits with CSF SAA Results** | | | | | **Characteristics at the Time of First SAA Result** | | | |
| --- | --- | --- | --- | --- | --- | --- | --- | --- | --- | --- |
| **Cohort** | **Subgroup** | **N with > 1 result** | **N first visit pos./type I** | **N first visit negative** | **N first visit type II** | **N first visit inconclusive** | **Age, mean (SD)** | **Sex,**  **% male** | **Duration, mean (SD)** | **MDS-UPDRS Part III OFF**, mean (SD)** |
| **PD** | **Total** | **633** | **493 (77.9%)** | **126 (19.9%)** | **9 (1.4%)** | **5 (0.8%)** | **62.6 (9.1)** | **399 (63.0%)** | **1.3 (1.9)** | **21.8 (9.7)** |
|  | sPD | 444 | 368 (82.9%) | 65 (14.6%) | 7 (1.6%) | 4 (0.9%) | 62.6 (9.0) | 300 (67.6%) | 0.6 (0.6) | 21.6 (9.3) |
|  | *LRRK2* PD | 125 | 76 (60.8%) | 48 (38.4%) | 1 (0.8%) | 0 | 63.7 (8.5) | 65 (52.0%) | 2.7 (2.1) | 21.5 (10.6) |
|  | *GBA* PD | 42 | 35 (83.3%) | 5 (11.9%) | 1 (2.4%) | 1 (2.4%) | 62.3 (9.6) | 22 (52.4%) | 3.0 (2.2) | 25.9 (11.3) |
|  | Other Genetic PD | 22 | 14 (63. 6%) | 8 (36.4%) | 0 | 0 | 57.1 (11.2) | 12 (54.5%) | 3.9 (5.8) | 19.1 (8.6) |
| **Prodromal** | **Total*** | **563** | **307 (54.5%)** | **247 (43.9%)** | **6 (1.1%)** | **3 (0.5%)** | **67.2 (6.0)** | **274 (48.7%)** | **-** | **4.6 (5.1)** |
|  | PSG+ iRBD & Hyposmia | 63 | 60 (95.2%) | 3 (4.8%) | 0 | 0 | 68.8 (5.7) | 49 (77.8%) | - | 4.4 (3.8) |
|  | PSG+ iRBD & Normosmia | 38 | 12 (31.6%) | 25 (65.8%) | 1 (2.6%) | 0 | 64.9 (6.8) | 32 (84.2%) | - | 3.7 (3.6) |
|  | Self-Reported DEB & Hyposmia | 139 | 102 (73.4%) | 34 (24.5%) | 3 (2.2%) | 0 | 68.4 (5.4) | 89 (64.0%) | - | 5.3 (5.5) |
|  | Hyposmia Only | 261 | 121 (46.4%) | 137 (52.5%) | 1 (0.4%) | 2 (0.8%) | 67.1 (5.0) | 75 (28.7%) | - | 4.2 (5.2) |
|  | Genetic NMCs | 58 | 10 (17.2%) | 46 (79.3%) | 1 (1.7%) | 1 (1.7%) | 64.5 (9.1) | 26 (44.8%) | - | 4.9 (5.4) |
| **HC** | **Total** | **42** | 18 (42.9%) | 23 (54.8%) | 0 | 1 (2.4%) | **66.7 (10.6)** | **28 (66.7%)** | **-** | **1.8 (2.2)** |

*Four participants in the “Other Prodromal” subgroup are included in the “Total” group but not listed separately. Of these, two had pos./Type I SAA and two had negative SAA at their first visit.

**49 participants are missing MDS-UPDRS III scores: 2 sPD, 23 *LRRK2* PD, 5 *GBA* PD, 3 other Genetic PD, 1 PSG+ iRBD & Hyposmia, 3 Self-Reported DEB & Hyposmia, 9 Hyposmia Only, and 3 Genetic NMCs

The “Other Genetic PD” group includes 17 PARKIN, 3 SNCA, and 2 LRRK2 + GBA carrier participants.

The “Genetic NMC” group includes 25 LRRK2, 21 GBA, 6 PARKIN, 3 SNCA, 2 LRRK2 + GBA, and 1 PARK7 carrier participants.

Abbreviations: sPD: sporadic PD. LRRK2: Leucine rich repeat kinase 2. GBA: glucosylceramidase beta 1. PSG: polysomnogram. iRBD: idiopathic REM sleep behavior disorder. DEB: dream enactment behavior. NMC: non-manifesting carrier. HC: healthy control

Supplementary Table 2. Longitudinal consistency of CSF aSyn-SAA results among those with N > 1 visits with clinical characteristics

|  | | | | | **Participants who remain pos./type I longitudinally** | | | | | | **Time interval between aSyn-SAA results**** | | | **Characteristics at First aSyn-SAA Visit** | | | | | | | | |
| --- | --- | --- | --- | --- | --- | --- | --- | --- | --- | --- | --- | --- | --- | --- | --- | --- | --- | --- | --- | --- | --- | --- |
| **Cohort** | **Subgroup** | | | **N first visit type I** | **N** | | **Percent (95% CL*)** | | | **Median # visits** **(min, max)** | | **Median duration (years)** **between first and last CSF aSyn-SAA visit (min, max)** | | | | **Age, mean (SD)** | | **Sex, % male** | | **Duration (years) from Dx, mean (SD)** | **MDS-UPDRS Part III OFF***, mean (SD)** | |
| **PD** | **Total** | | | **493** | **474** | | **96% (94%, 98%)** | | | **2.0 (2.0, 5.0)** | | **1.1 (0.4, 9.0)** | | | | **61.8 (8.8)** | | **322 (65%)** | | **1.2 (1.5)** | **22.0 (9.8)** | |
|  | sPD | | | 368 | 356 | | 97% (94%, 98%) | | | 2.0 (2.0, 4.0) | | 1.1 (0.4, 7.4) | | | | 62.0 (8.9) | | 250 (68%) | | 0.6 (0.6) | 21.4 (9.3) | |
|  | *LRRK2* PD | | | 76 | 74 | | 97% (91%, 99%) | | | 2.0 (2.0, 4.0) | | 1.0 (0.9, 5.0) | | | | 60.9 (8.3) | | 46 (61%) | | 2.8 (2.1) | 23.6 (11.6) | |
|  | *GBA* PD | | | 35 | 31 | | 89% (74%, 95%) | | | 2.0 (2.0, 4.0) | | 1.1 (0.9, 9.0) | | | | 63.1 (8.5) | | 19 (54%) | | 3.3 (2.2) | 27.5 (10.7) | |
|  | Other Genetic PD | | | 14 | 13 | | 93% (69%, 99%) | | | 2.0 (2.0, 5.0) | | 1.0 (1.0, 5.5) | | | | 59.5 (10.1) | | 7 (50%) | | 1.5 (2.0) | 20.3 (9.8) | |
| **Prodromal** | **Total****** | | | **307** | **303** | | **99% (97%, 99%)** | | | **2.0 (2.0, 7.0)** | | **2.1 (0.4, 5.2)** | | | | **68.5 (5.7)** | | **186 (61%)** | | **-** | **4.8 (5.2)** | |
|  | PSG+ iRBD & Hyposmia | | | 60 | 60 | | 100% (94%, 100%) | | | 2.0 (2.0, 7.0) | | 2.1 (0.4, 5.1) | | | | 68.8 (5.8) | | 46 (77%) | | - | 4.4 (3.8) | |
|  | PSG+ iRBD & Normosmia | | | 12 | 11 | | 92% (65%, 99%) | | | 2.0 (2.0, 5.0) | | 2.1 (1.9, 3.0) | | | | 67.4 (6.8) | | 12 (100%) | | - | 3.6 (2.7) | |
|  | Self-Reported DEB & Hyposmia | | | 102 | 101 | | 99% (95%, 100%) | | | 2.0 (2.0, 2.0) | | 2.0 (0.8, 2.4) | | | | 68.9 (5.6) | | 70 (69%) | | - | 5.1 (5.0) | |
|  | Hyposmia Only | | | 121 | 119 | | 98% (94%, 100%) | | | 2.0 (2.0, 7.0) | | 2.1 (0.5, 5.2) | | | | 68.2 (5.1) | | 51 (42%) | | - | 4.9 (6.3) | |
|  | Genetic NMCs | | | 10 | 10 | | 100% (72%, 100%) | | | 2.0 (2.0, 6.0) | | 2.1 (0.7, 3.7) | | | | 65.4 (11.2) | | 6 (60%) | | - | 3.4 (2.2) | |
| **HC** | **Total** | | | **18** | **16** | | **89% (67%, 97%)** | | | **2.0 (2.0, 4.0)** | | **2.5 (1.0, 10.8)** | | | | **68.2 (8.0)** | | **14 (78%)** | | - | **1.9 (2.7)** | |
| *95% Wilson Confidence Intervals.  **The “Hyposmia Only” subgroup has one participant with missing time interval information.  ***31 participants are missing MDS-UPDRS III scores: 1 sPD, 18 LRRK2 PD, 4 GBA PD, 1 other Genetic PD, 1 PSG+ iRBD & Hyposmia, 2 Self-Reported DEB & Hyposmia, and 4 Hyposmia Only.  ****Two participants in the “Other Prodromal” subgroup are included in the “Total” group but not listed separately. Both participants remained pos./Type I longitudinally. | | | | | | | | | | | | | | | | | | | | | | |
|  | | | | | | **Participants who remain negative longitudinally** | | | **Time interval between CSF aSyn-SAA results**** | | | | | | **Characteristics at First aSyn-SAA Visit** | | | | | | | |
| **Cohort** | | **Subgroup** | **N first visit negative** | | | **N** | | **Percent (95% CL*)** | **Median # visits** **(min, max)** | | | | **Median duration (years)** **between first and last aSyn-SAA visit** **(min, max)** | | **Age, mean (SD)** | | **Sex, % male** | | **Duration (years) from Dx, mean (SD)** | | | **MDS-UPDRS Part III OFF***, mean (SD)** |
| **PD** | | **Total** | **126** | | | **116** | | **92% (86%, 96%)** | **2.0 (2.0, 3.0)** | | | | **1.1 (0.9, 9.4)** | | **66.0 (9.3)** | | **70 (56%)** | | **1.9 (3.0)** | | | **20.1 (8.6)** |
|  | | sPD | 65 | | | 56 | | 86% (76%, 93%) | 2.0 (2.0, 3.0) | | | | 1.1 (0.9, 9.4) | | 66.3 (9.3) | | 43 (66%) | | 0.7 (0.5) | | | 21.9 (8.6) |
|  | | *LRRK2* PD | 48 | | | 48 | | 100% (93%, 100%) | 2.0 (2.0, 2.0) | | | | 1.0 (0.9, 7.0) | | 68.2 (6.9) | | 19 (40%) | | 2.6 (2.1) | | | 18.6 (8.7) |
|  | | *GBA* PD | 5 | | | 4 | | 80% | 2.0 (2.0, 2.0) | | | | 1.8 (0.9, 5.0) | | 60.8 | | 3 (60%) | | 1.8 | | | 15.0 |
|  | | Other Genetic PD | 8 | | | 8 | | 100% | 2.0 (2.0, 2.0) | | | | 1.0 (0.9, 3.2) | | 52.9 | | 5 (63%) | | 8.2 | | | 16.3 |
| **Prodromal** | | **Total****** | **247** | | | **234** | | **95% (91%, 97%)** | **2.0 (2.0, 6.0)** | | | | **2.0 (0.6, 8.9)** | | **65.8 (5.7)** | | **84 (34%)** | | - | | | **4.2 (4.6)** |
|  | | PSG+ iRBD & Normosmia | 25 | | | 25 | | 100% (87%, 100%) | 2.0 (2.0, 6.0) | | | | 2.0 (1.9, 7.2) | | 63.8 (6.8) | | 19 (76%) | | - | | | 3.7 (4.0) |
|  | | Self-Reported DEB & Hyposmia | 34 | | | 32 | | 94% (81%, 98%) | 2.0 (2.0, 3.0) | | | | 2.0 (0.6, 2.3) | | 66.8 (4.9) | | 17 (50%) | | - | | | 5.2 (5.3) |
|  | | Hyposmia Only | 137 | | | 132 | | 96% (92%, 98%) | 2.0 (2.0, 3.0) | | | | 2.0 (0.8, 2.5) | | 66.1 (4.7) | | 24 (18%) | | - | | | 3.6 (4.0) |
|  | | Genetic NMCs | 46 | | | 41 | | 89% (77%, 95%) | 2.0 (2.0, 6.0) | | | | 4.0 (1.0, 8.9) | | 65.1 (8.0) | | 19 (41%) | | - | | | 5.3 (5.9) |
| **HC** | | **Total** | **23** | | | **20** | | **87% (68%, 95%)** | **2.0 (2.0, 5.0)** | | | | **2.1 (0.8, 11.4)** | | **65.4 (12.4)** | | **14 (61%)** | | - | | | **1.5 (1.8)** |
| *95% Wilson Confidence Intervals.  **The “Hyposmia Only” subgroup has one participant with missing time interval information.  ***17 participants are missing MDS-UPDRS III scores: 1 sPD, 5 LRRK2 PD, 1 GBA PD, 2 other Genetic PD, 1 Self-Reported DEB & Hyposmia, 5 Hyposmia Only, and 2 Genetic NMCs.  ****Three participants in the “PSG+ iRBD & Hyposmia” subgroup and two participants in the “Other Prodromal” subgroup are included in the “Total” group but not listed separately. 67% of the “PSG+ iRBD & Hyposmia group” and 100% of the “Other Prodromal” group remained negative longitudinally. | | | | | | | | | | | | | | | | | | | | | | |

Supplementary Table 3a. CSF aSyn-SAA longitudinal consistency among prodromal-PD participants with an initial type I positive result – capped at Year 2

|  | | | **Participants who remain pos./Type I through Year 2** | |  | **Characteristics at First SAA Visit** | | |
| --- | --- | --- | --- | --- | --- | --- | --- | --- |
| **Cohort** | **Subgroup** | **N first visit type I** | **N** | **Percent (95% CL*)** | **Median # visits (min, max)** | **Age, mean (SD)** | **Sex, % male** | **MDS-UPDRS Part III OFF**, mean (SD)** |
| Prodromal | Total Prodromal*** | 285 | 281 | 99% (96%, 99%) | 2.0 (2.0, 4.0) | 68.5 (5.5) | 175 (61%) | 4.7 (5.2) |
|  | PSG+ iRBD & Hyposmia | 53 | 53 | 100% (93%, 100%) | 2.0 (2.0, 4.0) | 68.9 (6.1) | 40 (75%) | 4.3 (3.7) |
|  | PSG+ iRBD & Normosmia | 12 | 11 | 92% (65%, 99%) | 2.0 (2.0, 4.0) | 67.4 (6.8) | 12 (100%) | 3.6 (2.7) |
|  | Self-Reported DEB & Hyposmia | 100 | 99 | 99% (95%, 100%) | 2.0 (2.0, 2.0) | 68.9 (5.6) | 69 (69%) | 5.1 (5.0) |
|  | Hyposmia Only | 112 | 110 | 98% (94%, 100%) | 2.0 (2.0, 4.0) | 67.9 (4.9) | 47 (42%) | 4.8 (6.2) |
|  | Genetic NMCs | 7 | 7 | 100% | 2.0 (2.0, 4.0) | 70.3 | 6 (86%) | 3.0 |
| *95% Wilson Confidence Intervals.  **7 participants are missing MDS-UPDRS III scores: 1 PSG+ iRBD & Hyposmia, 2 Self-Reported DEB & Hyposmia, and 4 Hyposmia Only  ***One participant in the “Other Prodromal” subgroup included in the “Total” group but not listed separately.  *The Genetic NMC group includes 3 GBA, 1 PARK7, and 3 PARKIN carriers.* | | | | | | | | |

Supplementary Table 3b. CSF aSyn0-SAA longitudinal consistency among those with an initial negative result – capped at Year 2

|  | | | **Participants who remain negative through Year 2** | |  | **Characteristics at First SAA Visit** | | |
| --- | --- | --- | --- | --- | --- | --- | --- | --- |
| **Cohort** | **Subgroup** | **N first visit negative** | **N** | **Percent (95% CL*)** | **Median # visits (min, max)** | **Age, mean (SD)** | **Sex, % male** | **MDS-UPDRS Part III OFF**, mean (SD)** |
| Prodromal | Total Prodromal*** | 182 | 175 | 96% (92%, 98%) | 2.0 (2.0, 4.0) | 66.1 (5.6) | 61 (34%) | 3.9 (4.6) |
|  | PSG+ iRBD & Normosmia | 24 | 24 | 100% (86%, 100%) | 2.0 (2.0, 4.0) | 63.9 (6.9) | 18 (75%) | 3.5 (4.0) |
|  | Self-Reported DEB & Hyposmia | 23 | 22 | 96% (79%, 99%) | 2.0 (2.0, 3.0) | 66.8 (4.3) | 12 (52%) | 5.8 (6.0) |
|  | Hyposmia Only | 115 | 110 | 96% (90%, 98%) | 2.0 (2.0, 3.0) | 66.3 (4.9) | 24 (21%) | 3.4 (3.9) |
|  | Genetic NMCs | 16 | 16 | 100% (81%, 100%) | 2.0 (2.0, 3.0) | 66.9 (8.9) | 3 (19%) | 5.5 (6.6) |
| *95% Wilson Confidence Intervals.  **5 participants are missing MDS-UPDRS III scores: 4 Hyposmia Only, and 1 Genetic NMC  ***There is 1 participant in the “PSG+ iRBD & Hyposmia” subgroup and 2 participants in the “Other Prodromal” subgroup included in the “Total” group but not listed separately.  *The Genetic NMC group includes 7 GBA, 4 LRRK2, 1 LRRK2+GBA, 3 PARKIN, and 1 SNCA carrier.* | | | | | | | | |

Abbreviations: sPD: sporadic PD. LRRK2: Leucine rich repeat kinase 2. GBA: glucosylceramidase beta 1. PSG: polysomnogram. iRBD: idiopathic REM sleep behavior disorder. DEB: dream enactment behavior. NMC: non-manifesting carrier. HC: healthy control

**PPMI STUDY COMMITTEES, CORES, AND COLLABORATORS**

**PPMI EXECUTIVE STEERING COMMITTEE**

Kenneth Marek, MD 1 (Principal Investigator); Tanya Simuni, MD 2 ; Andrew Siderowf, MD 3 ; Caroline Tanner, MD 4 ; Thomas F

Tropea, DO 1 ; Tatiana Foroud, PhD 5 ; Lana Chahine, MD 6 ; Brit Mollenhauer, MD 7 ; Kalpana Merchant, MD 2 ; Douglas Galasko,

MD 8; Christopher Coffey, PhD 9 ; Kathleen Poston, MD 10 ; Roseanne Dobkin, PhD 11 ; Ethan Brown, MD 4 ; Roy Alcalay, MD 12 ; Dan

Weintraub, MD 3 ; Emily Flagg, BA 1 ; Kimberly Fabrizio, BA 1

**PPMI STEERING COMMITTEE**

Susan Bressman, MD 13 ; Cornelis Blauwendraat, PhD 14 ; Paola Casalin, PhD 15 ; Sonya Dumanis, PhD 14 ; Raymond James, RN 16 ;

Karl Kieburtz, MD 17 ; Sneha Mantri, MS 18 ; Werner Poewe, MD 19 ; Michael Schwarzschild, MD20 ; John Seibyl1 , MD; David

Standaert, PhD 21 ; Duygu Tosun-Turgut, PhD 4

**MICHAEL J. FOX FOUNDATION**

Sohini Chowdhury, MA 22 ; Jamie Eberling, PhD 22 ; Mark Frasier, PhD 22 ; Leslie Kirsch, EdD 22 ; Katie Kopil, PhD 22 ; Maggie Kuhl,

BA 22 ; Alyssa O’Grady, BA 22 ; Todd Sherer, PhD 22 ; Tawny Willson, MBS22

**PPMI STUDY CORES**

Project Management Core: Emily Flagg, BA 1

Site Management Core: Tanya Simuni, MD 2 ; Bridget McMahon, BS 1

Data Strategy and Technical Operations: Craig Stanley, PhD 1 ; Kim Fabrizio, BA 1

Data Management Core: Dixie Ecklund, MBA 9 , MSN; Christine Kohnen, PhD 9

Screening Core: Tatiana Foroud, PhD 5 ; Laura Heathers, BA 5 ; Christopher Hobbick, BSCE5 ; Gena Antonopoulos, BSN 5

Imaging Core: John Seibyl, MD 1 ; Kathleen Poston, MD 10

Statistics Core: Christopher Coffey, PhD 9 ; Chelsea Caspell-Garcia, MS 9 ; Michael Brumm, MS 9

Bioinformatics Core: Arthur Toga, PhD 23 ; Karen Crawford, MLIS 23

Biorepository Core: Tatiana Foroud, PhD 5 ; Jan Hamer, BS5

Biologics Review Committee: Brit Mollenhauer, MD 7 ; Doug Galasko, MD 8 ; Kalpana Merchant, MD 2

Genetics Core: Andrew Singleton, PhD 24

Pathology Core: Tatiana Foroud, PhD 5 ; Dirk Keene, MD 5

Found: Caroline Tanner, MD 4 ; Ethan Brown, MD 4

PPMI Online: Carlie Tanner, MD 4 ; Ethan Brown, MD 4 ; Lana Chahine, MD 6 ; Roseann Dobkin, PhD 11 ; Monica Korell, MPH 4

**PPMI SITE INVESTIGATORS**

Neha Prakash MD1 ; Tanya Simuni, MD2 ; Nabila Dahodwala MD3 ; Caroline Tanner, MD4 ; Lana Chahine MD6 ; Brit Mollenhauer MD7 ; Sebastian

Schade MD7 ; Douglas Galasko, MD 8 ; Anat Mirelman PhD12 ; Roy Alcalay MD12 ;Katherine Leaver MD13 ; Marie Saint-Hilaire MD16 ; Ruth

Schneider MD17 ; Christopher Tarolli MD17 ; Werner Poewe, MD19 ; Aleksandar Videnovic MD20 ; David Standaert PhD21 ; Marissa Dean, MD21 ;

Sonja Jonsdottir PhD25 ; Rejko Krueger MD25 ; Claire Pauly PhD25 ; Stewart Factor DO 26 ; Penelope Hogarth MD26 ; Robert Hauser MD28 ; Amy

Amara PhD29 ; Michelle Fullard MD29 ; Cyrus Zabetian MD30 ; Hubert Fernandez MD31 ; Kathrin Brockmann MD32 ; Isabel Wurster PhD32 ; Yen

Tai PhD33 ; Paolo Barone PhD34 ; Marina Picillo MD34 ; Stuart Isaacson MD35 ; Alberto Espay MD36 ; Eduardo Tolosa PhD37 ; Javier Ruiz Martinez

PhD38 ; Leonidas Stefanis PhD39 ; Kelvin Chou MD40 ; Lorraine Kalia MD41 ; Connie Marras PhD41 ; David Grimes MD42 ; Tiago Mestre PhD42 ;

Rajesh Pahwa MD43 ; Mark Lew MD44 ; Holly Shill MD45 ; Shyamal Mehta MD46 ; Giulietta Riboldi MD47 ; Nikolaus McFarland PhD48 ; Ron

Postuma MD49 ; Zoltan Mari MD50 ; David Ledingham MD51 ; Nicola Pavese PhD51 ; Michele Hu PhD52 ; Norbert Brueggemann MD53 , ; Christine

Klein MD53 ; Bastiaan Bloem PhD54 ; Cristina Simonet PhD55 ; Alastair Noyce PhD55 ; Anette Janzen PhD56 ; David Pedrosa MD56 ; Wolfgang

Oertel PhD56 ; Njideka Okubadejo MD57 David Shprecher DO 58 ; Arjun Tarakad MD59 ; Emile Moukheiber MD60

**PPMI SITE COORDINATORS**

Joy Antala¹; Carla Aranda²; Karen Williams²; Sophia Melton²; Karina Benson²; Ashwini Ramachandran³; Danielle Potts³; Grace LaMoure³;

Ritikha Vengadesh3 ; Ryan Manzler³; Jaime Heller⁴; Primi Ranola⁴; Farah Kausar4 ; Sherri Mosovsky⁶; Diana Willeke⁷; Elizabeth Kalinkara-

Gomez⁷; Janelle Rodriguez⁸; Nobuko Kemmotsu 8 ; May Eshel¹²; Deborah Raymond¹³; Abigail Desrosiers16 ; Raymond James¹⁶; Lauren

Jackson¹⁷; Iris Egner¹⁹; Wesley Schlett²⁰; Courtney Blair²¹; Lauren Ruffrage²¹; Berenice Sevilla²⁵; Barbara Sommerfeld²⁶; Dustin Le²⁷; Erica

Botting²⁸; Gabriella Mazur²⁸; Daniele Derlein²⁹; Evan Doll²⁹; Ying Liu²⁹; Ciera Cobb³⁰; Olivia Masiewicz³⁰; Jennifer Mule³¹; Michael Morsillo³¹;

Ella Hilt³²; Aldazier Jakiran 33 ; Dominga Valentino34 ; Lisbeth Pennente 35 ; Bobbie Stubbeman³⁶; Alicia Garrido³⁷; Valeria Ravasi³⁷; Ioana

Croitoru38 ; Christos Koros³⁹; Nikolas Papagiannakis³⁹; Frank Ferrari⁴⁰; Mengyu Zheng⁴¹; Shawna Reddie⁴²; Alicia Alejandra⁴³; Andrea Gray⁴³;

Alejandra Valenzuela 44 ; Caitlin Goodman 45 ; Sara Dresler⁴⁶; Neil Santos46 ; Fahrial Esha⁴⁷; Kyle Rizer⁴⁸; Nadine Zablith⁴⁹; Liliana

Dumitrescu⁵⁰; Debra Galley⁵¹; Victoria Kate Foster⁵¹; Jamil Razzaque⁵²; Madita Grümmer⁵³; Yara Krasowski⁵⁴; Natalie Donkor⁵⁵; Elisabeth

Sittig⁵⁶; Oluwadamilola Ojo⁵⁷; Kelly Clark⁵⁸; Rory Mahabir⁵⁹; Kori Ribb⁶⁰; Shamera Willoughby⁶⁰

June 2025

**INSTITUTIONS AND AFFILIATIONS**

1. Institute for Neurodegenerative Disorders; New Haven, CT, USA

2. Northwestern University; Evanston, IL, USA

3. University of Pennsylvania; Philadelphia, PA, USA

4. University of California, San Francisco; San Francisco, CA, USA

5. Indiana University; Indianapolis, IN, USA

6. University of Pittsburgh; Pittsburgh, PA, USA

7. Paracelsus-Elena Klinik; Kassel, Germany

8. University of California, San Diego, San Diego, CA, USA

9. University of Iowa; Iowa City, IA, USA

10. Stanford University; Stanford, CA, USA

11. Rutgers University; New Brunswick, NJ, USA

12. Tel Aviv Sourasky Medical Center; Tel Aviv, Israel

13. Mount Sinai Beth Israel; New York, NY, USA

14. Coalition for Aligning Science; Chevy Chase, MD, USA

15. BioRep; Milan, Italy

16. Boston University School of Medicine; Boston, MA, USA

17. University of Rochester; Rochester, NY, USA

18. Duke University; Durham, NC, USA

19. University of Innsbruck; Innsbruck, Austria

20. Massachusetts General Hospital; Boston, MA, USA

21. University of Alabama at Birmingham; Birmingham, AL, USA

22. The Michael J. Fox Foundation for Parkinson’s Research; New York, NY, USA

23. Laboratory of Neuroimaging (LONI), USC; Los Angeles, CA, USA

24. National Institute on Aging, NIH; Bethesda, MD, USA

25. University of Luxembourg; Esch-sur-Alzette, Luxembourg

26. Emory University; Atlanta, GA, USA

27. Oregon Health and Science University; Portland, OR, USA

28. University of South Florida; Tampa, FL, USA

29. University of Colorado; Aurora, CO, USA

30. VA Puget Sound Health System; Seattle, WA, USA

31. Cleveland Clinic; Cleveland, OH, USA

32. University of Tubingen; Tubingen, Germany

33. Imperial College of London; London, UK

34. University of Salerno; Salerno, Italy

35. Parkinson’s Disease and Movement Disorders Center; Boca Raton, FL, USA

36. University of Cincinnati; Cincinnati, OH, USA

37. Hospital Clinic of Barcelona; Barcelona, Spain

38. Hospital Universitario Donostia; San Sebastian, Spain

39. University of Athens; Athens, Greece

40. University of Michigan; Ann Arbor, MI, USA

41. Toronto Western Hospital; Toronto, Canada

42. The Ottawa Hospital; Ottawa, Canada

43. University of Kansas Medical Center; Kansas City, KS, USA

44. Keck School of Medicine of the University of Southern California; Los Angeles, CA, USA

45. Barrow Neurological Institute; Phoenix, AZ, USA

46. Mayo Clinic Arizona; Scottsdale, AZ, USA

47. NYU Langone Medical Center; New York, NY, USA

48. University of Florida; Gainesville, FL, USA

49. Montreal Neurological Institute and Hospital/McGill; Montreal, QC, Canada

50. Cleveland Clinic-Las Vegas Lou Ruvo Center for Brain Health; Las Vegas, NV, USA

51. Clinical Ageing Research Unit; Newcastle, UK

52. John Radcliffe Hospital Oxford and Oxford University; Oxford, UK

53. University of Luebeck; Luebeck, Germany

54. Radboud University; Nijmegen, Netherlands

55. Queen Mary University of London; London, UK

56. Philipps-University Marburg; Marburg, Germany

57. University of Lagos; Lagos, Nigeria

58. Banner Sun Health Research Institute; Sun City, AZ, USA

59. Baylor College of Medicine; Houston, TX, USA

60. Johns Hopkins University; Baltimore, MD, USA

**PPMI STUDY FUNDING PARTNER STATEMENT**

PPMI – a public-private partnership – is funded by the Michael J. Fox Foundation for Parkinson’s Research and funding partners, including 4D Pharma, Abbvie,

AcureX, Allergan, Amathus Therapeutics, Aligning Science Across Parkinson's, AskBio, Avid Radiopharmaceuticals, BIAL, BioArctic, Biogen, Biohaven,

BioLegend, BlueRock Therapeutics, Bristol-Myers Squibb, Calico Labs, Capsida Biotherapeutics, Celgene, Cerevel Therapeutics, Coave Therapeutics, DaCapo

Brainscience, Denali, Edmond J. Safra Foundation, Eli Lilly, Gain Therapeutics, GE HealthCare, Genentech, GSK, Golub Capital, Handl Therapeutics, Insitro,

Jazz Pharmaceuticals, Johnson & Johnson Innovative Medicine, Lundbeck, Merck, Meso Scale Discovery, Mission Therapeutics, Neurocrine Biosciences,

Neuron23, Neuropore, Pfizer, Piramal, Prevail Therapeutics, Roche, Sanofi, Servier, Sun Pharma Advanced Research Company, Takeda, Teva, UCB, Vanqua Bio,

Verily, Voyager Therapeutics, the Weston Family Foundation and Yumanity Therapeutics.
